# PNEUMOCOCCAL CONJUGATE VACCINE SCHEDULES SHAPE SEROTYPE-SPECIFIC MEMORY B-CELL RESPONSES

**DOI:** 10.64898/2026.11.09.26367657

**Authors:** Hugh Elliott Brown, Thanh V Phan, Zheng Quan Toh, Beth Temple, Cattram Duong Nguyen, Kien Vientrung, Hoang Van Anh Nguyen, Nhung Vu Pham Hong, Vo Thi Trang Dai, Kathryn Bright, Hau Phuc Tran, Rachel Ann Higgins, Nadia Mazarakis, Skyy Luu, Leanne Quah, Daniel G Pellicci, Thuong Vu Nguyen, Jeremy Anderson, Kim Mulholland, Paul Vincent Licciardi

## Abstract

Reduced-dose pneumococcal conjugate vaccine (PCV) schedules (‘1+1’; one primary and one booster) are now endorsed by the World Health Organisation under specific circumstances. Serotype-specific memory B cells (B_MEM_) are a marker of long-term protection but data from infant reduced-dose trials remain limited. As part of a randomised controlled trial in Vietnamese infants, we characterised serotype-specific B_MEM_ responses in randomised subsets (N=50 per vaccine group per timepoint) following 0+1 (at 12 months of age) or 1+1 (2 and 12 months of age) schedules of PCV10 or PCV13, using a multimer spectral flow cytometry approach. Blood samples were collected at 12 months of age (baseline) and at day 7 and day 28 post-vaccination.

A one or two dose schedule of either PCV induced robust serotype-specific B_MEM_ responses at day 7 and day 28 for most serotypes. PCV13 generated earlier and higher B_MEM_ responses than PCV10, while 1+1 schedules for either PCV produced higher B_MEM_ responses than the 0+1 schedules at day 7. By day 28, these differences between the 0+1 and 1+1 schedules were no longer apparent. Notably, B_MEM_ responses for serotypes 6B and 23F were detectable even in infants who did not achieve protective levels of serotype-specific IgG ≥0.35 µg/mL. PCV13 generated higher plasmablast and plasma cell responses by day 7, and higher cTfh responses by day 28, compared to PCV10.

These findings show that 1+1 schedules of PCV10 or PCV13 induce robust B_MEM_ responses, with important implications for the evaluation of PCVs and understanding long-term protection.

## Introduction

*Streptococcus pneumoniae* (pneumococcus) is a leading cause of morbidity and mortality globally in children under 5 years of age^1,2^, mostly in low-and middle-income countries (LMICs) ^3,4^. Over 100 pneumococcal serotypes have been identified based on their capsular polysaccharide structure. Current pneumococcal conjugate vaccine (PCV) formulations licensed for use in infants include the 10-valent vaccines (Synflorix, GSK and Pneumosil, Serum Institute of India, SII), a 13-valent vaccine (Prevenar 13, Pfizer) ^5^, a 15-valent vaccine (Vaxneuvance, Merck), and a 20-valent vaccine (Prevenar 20, Pfizer).

The introduction of PCV into infant immunisation programmes has substantially reduced the incidence of pneumococcal disease, although pneumococcal carriage and disease caused by non-vaccine serotypes (serotype replacement) remains a significant issue ^3^. The World Health Organisation (WHO) recommends infant PCV schedules be delivered as a three-dose schedule either as two primary doses followed by a booster (2+1) or as a three-dose primary series without a booster (3+0). In the 2025 WHO PCV position statement, a single primary and single booster (1+1) schedule was added to this recommendation for use in settings with an established infant PCV programme . Currently, the United Kingdom is the only country to have implemented a 1+1 schedule.

PCVs generate protection through the induction of serotype-specific antibody responses and immunological memory. The gold-standard methods for evaluating PCV immunogenicity include serotype-specific IgG and opsonophagocytosis ^6^, which have established correlates of protection against invasive pneumococcal disease (IPD). As studies of IPD require very large sample sizes, pneumococcal carriage (the precursor to disease) is often used as a surrogate for clinical protection in addition to immunogenicity. However, unlike for IPD, there are no established correlates of protection against carriage. Protection against carriage has been demonstrated even when antibodies have waned ^7,8^, suggesting other immunological markers may play a role. For 1+1 schedules, this is important since it relies on protection against carriage (indirect protection) in the second year of life. Non-serological markers such as memory B cells (B_MEM_) may be an important marker of long-term protection following 1+1 PCV schedules, but there are no data from 1+1 randomised controlled trials in infants. Our earlier Bmem study in infants given a 1+1 schedule focused on a limited number of serotypes ^9^ and did not include detailed phenotypic data of vaccine-induced B cells and circulating follicular T helper cells (cTfh) which are important in memory formation and antibody responses.

We conducted a randomised controlled trial (RCT) to examine the immunogenicity and impact on carriage of 1+1 (2m and 12m) and 0+1 (12m) PCV10 or PCV13 schedules in Vietnamese infants. Primary outcome carriage and immunogenicity data have been published previously ^10^. In this study, we provide detailed characterisation of serotype-specific B_MEM_ responses to all vaccine serotypes as well as measurement of memory B cell and cTfh populations post-vaccination using a spectral flow cytometry approach following reduced-dose schedules in Vietnamese infants.

## Methods

### Study design and participant samples

The Vietnam Pneumococcal Trial II (VPT-II; NCT03098628) was a randomised controlled trial that enrolled 2,501 infants across three districts in Ho Chi Minh City, to evaluate the immunogenicity and effects on carriage of a 0+1 (given at 12 months) and 1+1 (given at 2 months and 12 months) schedule of PCV10 or PCV13 ^10,11^. Subsets of participants (n=50 per vaccine group per timepoint) were further randomised to provide peripheral blood mononuclear cells (PBMCs) and plasma at one of the following time points: 12 months of age (baseline), day 7 post-vaccination, or day 28 post-vaccination as part of an immunology sub-study (Fig. 1A) ^11^. Each timepoint therefore comprises samples from different, independently randomised individuals rather than repeat measures on the same infants.

**Figure 1.**
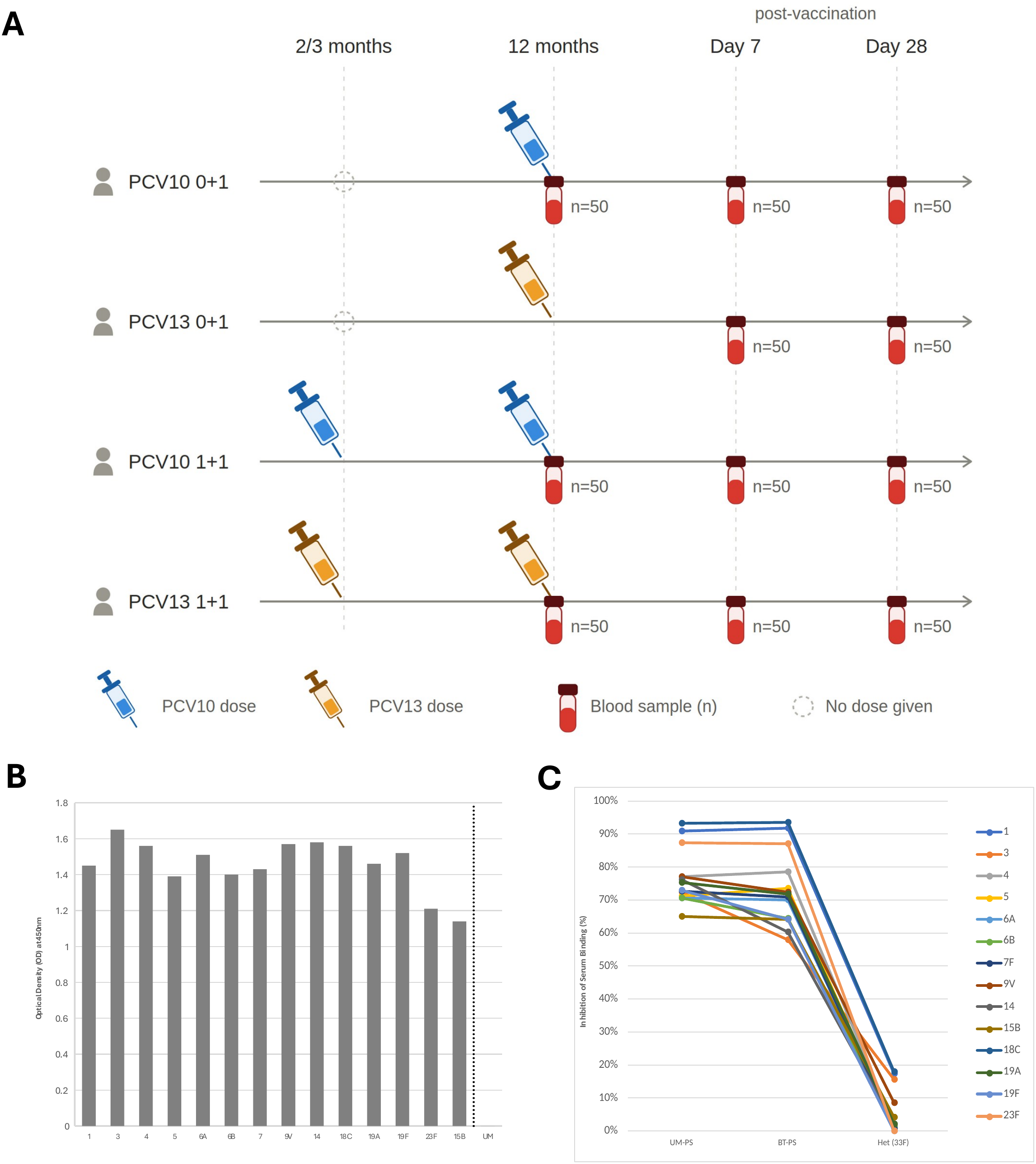
- Sample collection and validation **A**, Immunology sub-study timeline showing blood samples collected at 12 months, day 7 and day 28 post-vaccination. **B**, Biotin ELISA showing optical density (OD) at 450nm for biotinylated polysaccharides and unmodified (UM) polysaccharides. Each serotype was run in duplicate, and bars represent average of two replicate values after correcting for blanks. Unmodified polysaccharides were included as negative controls; all displayed an optical density of 0 therefore were represented as one bar. **C**, Competition assay showing proportion of serum inhibition by unmodified polysaccharides (UM-PS), biotinylated polysaccharides (BT-PS), and heterologous serotype 33F polysaccharide (het) for the 13 PCV serotypes and non-vaccine serotype 15B.

Ethical approval was obtained from the Royal Children’s Hospital Melbourne human research ethics committee and the Viet Nam Ministry of Health ethics committee.

### Serotype-specific IgG measurement

A modified WHO ELISA was used to determine serotype-specific IgG concentrations in serum samples at all timepoints ^12^. Full details are provided in Supplementary Methods.

### Multimer Preparation

Cyanylation and biotinylation of pneumococcal polysaccharides (PS) were performed as described previously ^13^, with minor modifications. This included the use of 4-Dimethylaminopyridine (DMAP) to buffer the cyanylation reaction ^14^ (Supplementary Fig. 1A, Supplementary Methods). Confirmation of biotinylation was performed by indirect ELISA (Supplementary Methods).

Multimers were made whereby biotinylated polysaccharides and conjugated streptavidin were diluted separately in PBS to a final concentration of 10µg/mL. The streptavidin working mix was added to the polysaccharide working mix in 10 steps at 1/10^th^ of total biotinylated PS volume with a 10 min incubation between each addition in a 96-well plate. After the final addition, plates were incubated for a further 30 min to ensure complete binding of biotinylated PS to streptavidin. A final concentration (12.5 μM) of D-biotin (Supplementary Table 1) dissolved in PBS was added to the multimer mix to block unbound binding sites of the streptavidin.

### Competition ELISA

The specificity of biotinylated polysaccharides was assessed using a competition-based WHO ELISA method. Briefly, biotinylated and unmodified polysaccharides were coated onto ELISA plates (5 µg/mL) and incubated overnight at RT. The WHO reference serum 007sp was used as it contains antibodies to all 23 serotypes in Pneumovax and was pre-adsorbed overnight with cell wall polysaccharide (10 µg/mL) and serotype 22F (5 µg/mL) following the WHO ELISA protocol. The 007sp serum was then pre-incubated for 3 h at 37°C with homologous or heterologous (serotype 33F) biotinylated or unmodified polysaccharide at a 50, 5, and 0.5 µg/mL before transfer to the coated ELISA plate. The ELISA was then developed and stopped according to the WHO ELISA method (Supplementary Methods).

### Spectral flow cytometry

Cryopreserved PBMCs were thawed at 37°C, diluted in RPMI1640 to a volume of 10 mL and centrifuged at 700g and 4°C for 3 min. Supernatant was discarded and the cells resuspended in RPMI. Cells were counted using a Bio-Rad TC20 automated cell counter using the trypan blue exclusion method. Following counting, cells were centrifuged at 700g for 3 min at 4°C, and resuspended in 100 µL RPMI, and transferred to a 96-well U-bottom cell culture plate. Plates were spun and supernatant removed. PBMCs were stained with fluorescently labelled monoclonal antibodies for flow analysis. PBMCs were incubated with 50 µL of stain 1 at RT for 20 min (Supplementary Table 2). After incubation, the cells were washed twice with 100 µL of flow buffer (PBS + 2% FBS) and resuspended in 50 µL of stain 2 (Supplementary Table 2). Following a 30 min incubation at 4°C, cells were washed twice with 100 µL of flow buffer. The cells were then resuspended in 100 µL of multimer staining mix (stain 3, Supplementary Table 2) and incubated at 4°C for 1 h. Following incubation, cells were washed twice with 100 µL flow buffer. Cells were then resuspended in 150 µL of flow buffer for acquisition on the Cytek Aurora.

Total B_MEM_ (CD3^-^CD19^+^CD27^+^) were selected from total B cells. B_MEM_ specific to each vaccine serotype as well as a control, non-vaccine serotype (15B) was identified using a barcoding strategy ^13^, with double positives for select fluorochrome pairs representing a single serotype-specific subset (Supplementary Fig. 1C). B cell subsets were gated from total B cells; B1-like/regulatory B cells (CD38^-/lo^CD20^+^CD1d^+^CD5^+^) and marginal zone-like B cells (CD38^-/lo^CD20^+^CD1d^+^CD5^-^). These innate-like populations of B cells were defined based on CD1d expression as a marker of splenic marginal zone b cells ^15^ and separated by the regulatory like/B1 fraction using CD5^16^. Plasmablasts were gated from total B_MEM_ gate (CD38^hi^CD20^-^). Plasma cells were defined as CD3^-^CD20^+^CD27^+^CD38^hi^CD138^+^. For circulating T follicular helper cells (cTfh), these were defined as CD3^+^CD19^-^CD4^+^CXCR5^+^PD-1^+^ and its subpopulations Tfh1 (CCR6^+^CXCR3^-^), Tfh2 (CCR6^-^CXCR3^-^), Tfh17 (CCR6^-^CXCR3^+^), Tfh1/17 (CCR6^+^CXCR3^+^) were also identified. ICOS, an activation marker used for Tfh, shows age-related inflammatory changes, and is only transiently expressed on activation ^17^. A full gating strategy is presented in Supplementary Figure 2.

### Statistical Analysis

Unmixed FCS files were uploaded to CellEngine, where cellular populations were manually gated (Supplementary Fig. 1) and serotype-specific B_MEM_ were quantified as a percentage of total B_MEM_. Only samples with >5,000 total B_MEM_ were analysed. Statistical analysis and visualisation were performed in R. Continuous immune-response data, including antibody concentrations, B_MEM_ frequencies, and frequencies of antigen-nonspecific cellular populations were summarised as medians; antibody data were additionally summarised as the proportion of participants achieving seroprotection (serotype-specific IgG ≥0.35 µg/mL). Immunoglobulin isotype and subclass distribution was assessed only in samples with ≥10 cells per serotype-specific population and reported as median proportions. Detailed statistical analysis is provided in the Supplementary Methods.

## Results

Demographics for this subset of participants have been reported previously ^9^. There were no major differences between the immunology subset and the overall trial population.

### Optimisation of multimer setup

Biotinylated pneumococcal polysaccharides (PS) were generated for all PCV serotypes (and serotype 15B) and validated by a biotin ELISA, showing a clear signal compared to native (unmodified) PS (Fig. 1B). Specificity of biotinylated PS was maintained with a similar proportion of homologous serotype inhibition to unmodified PS. No inhibition was observed when an unrelated, heterologous PS (serotype 33F) was used (Fig. 1C). A barcoding strategy utilising two fluorochrome combinations was used to identify each serotype (Supplementary Fig. 1B). An unvaccinated adult PBMC sample was included in each trial sample as a negative control (Supplementary Fig. 1D). Additionally, a non-vaccine type (serotype 15B) was included to demonstrate specificity.

### Comparison of serotype-specific IgG and B_MEM_ between 0+1 and 1+1 schedules

Figure 2 shows IgG and B_MEM_ responses for representative serotypes that are shared between PCV10 and PCV13 (1, 6B, 19F) as well as unique to PCV13 (serotype 6A). Responses for all PCV serotypes are available in Supplementary Fig. 4.

**Figure 2.**
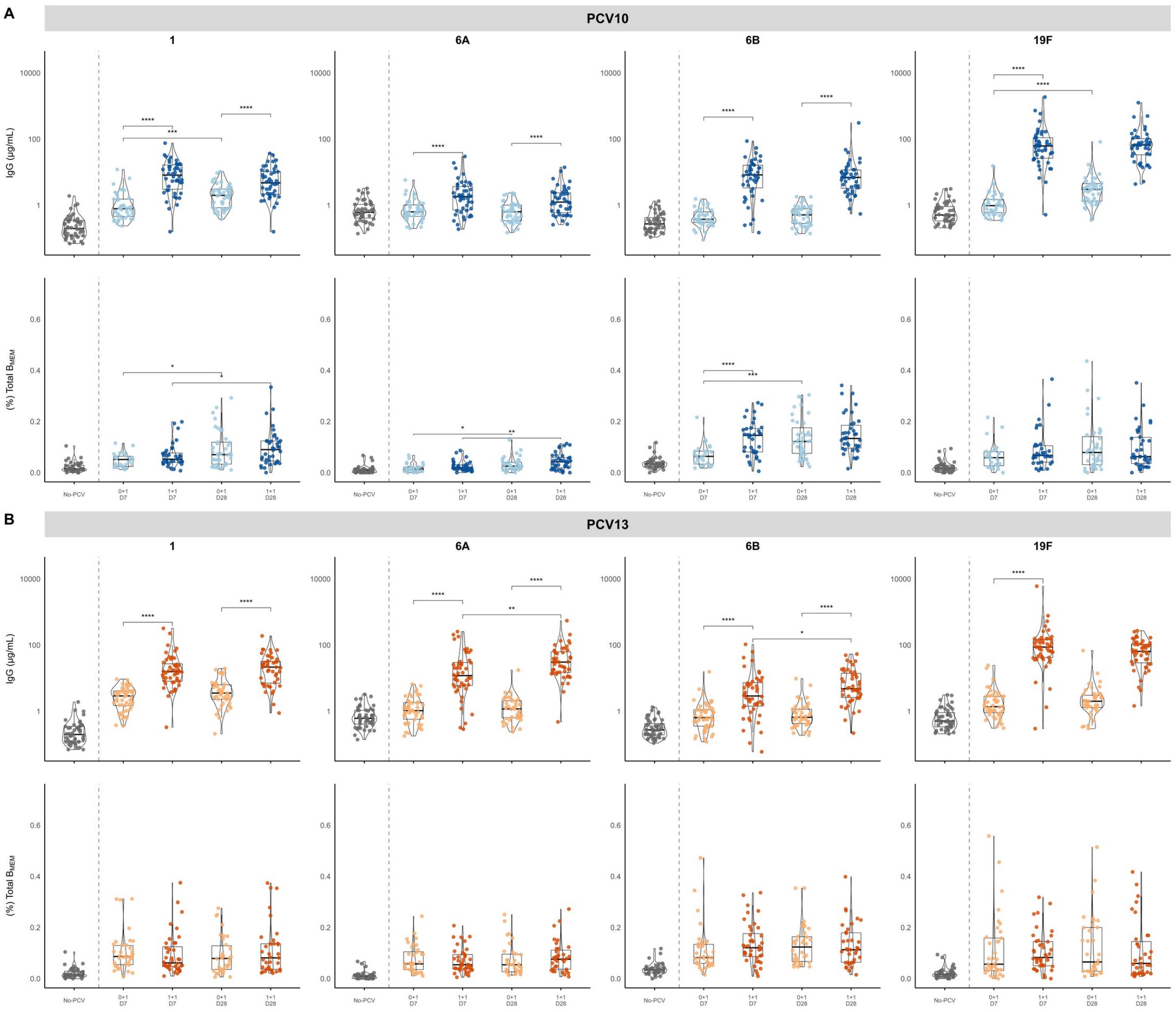
– Comparison of a 0+1 and 1+1 dosing schedule for PCV10 or PCV13 Serotype-specific B_MEM_ frequencies and serum IgG concentrations were measured at day 7 and day 28 post-vaccination following 0+1 and 1+1 dosing schedules. IgG data is shown for the subset of participants who were included in the immunology sub-study. Data are shown for PCV10 (**A**) and PCV13 (**B**). For the PCV10/PCV13 0+1 schedule (light blue/light orange), measurements are shown at 7-days and 28-days; for the PCV10/PCV13 1+1 schedule (dark blue/dark orange), measurements are shown at day 7 and day 28. A no-PCV comprising the PCV10 0+1 group at 12m (grey) is shown. Serotype-specific B_MEM_ are expressed as a proportion of total B_MEM_, and serotype-specific IgG concentrations are expressed as µg/mL. Box plots indicate the median and interquartile range, with violins showing the distribution within each group. Statistical significance was assessed using the Mann–Whitney test and is displayed for comparisons between 0+1 and 1+1 schedules at day 7 or day 28 post-vaccination, and between day 7 and day 28 time points within each dosing schedule. Significance is denoted as *p < 0.05, **p < 0.01, ***p < 0.005, ****p < 0.001.

For PCV10, IgG concentrations at 12m of age (no-PCV) were low for all serotypes (Fig. 2A, Supplementary Fig. 4A). In terms of seroprotection, in the PCV10 0+1 group, >90% of infants showed seroprotection (defined as serotype-specific IgG ≥ 0.35 µg/mL) for 4/10 serotypes at day 7 (serotypes 1, 5, 18C, and 19F; Supplementary Fig. 3A) and 7/10 serotypes at day 28 (the same four plus serotypes 4, 7F, and 9V; Supplementary Fig. 3B). In the PCV10 1+1 group, >90% of infants achieved seroprotection for all serotypes at both day 7 and day 28 (Supplementary Fig. 3). In the PCV13 0+1 group, >90% of infants achieved seroprotection at day 7 for 8/13 serotypes (except 6A, 6B, 7F, 18C, and 23F; Supplementary Fig. 3A), and 11/13 serotypes at day 28 (except 6B and 23F; Supplementary Fig. 3B). Seroprotection was >90% in the PCV13 1+1 group for all serotypes at both day 7 and day 28 (Supplementary Fig. 3).

Comparing IgG concentrations between the 0+1 and 1+1 PCV10 groups, these were higher in the 1+1 group for all serotypes at both time points, except for serotypes 3 and 18C at day 28, which were similar (Fig. 2A, Supplementary Fig. 4A). Comparing day 7 and day 28 in the PCV10 0+1 group, IgG concentrations were similar for all serotypes (except 1, 19F, 23F being higher at day 28). Comparing day 7 and day 28 time points in the PCV10 1+1 group, IgG concentrations were similar for all serotypes (Fig. 2A, Supplementary Fig. 4A).

Non-PCV10 serotypes did not increase from baseline at day 7 (3, 6A, 19A) or day 28 (3, 6A) in the PCV10 0+1 schedule, or for serotype 3 in the PCV10 1+1 group (Fig. 2A, Supplementary Fig. 4A). In the PCV13 0+1 group, IgG concentrations saw a modest increase for most serotypes from baseline at day 7 and a further increase at day 28. For the PCV13 1+1 group, most serotypes had a modest increase from baseline at day 7 and remained similar at day 28. For PCV13, IgG concentrations at baseline were low for all serotypes (Fig. 2B, Supplementary Fig. 4B). Comparing the 0+1 and 1+1 PCV13 groups, IgG concentrations were higher in the 1+1 group for all serotypes at day 7 and 12/13 serotypes at day 28 (except 7F; Fig. 2B, Supplementary Fig. 4B). In the 0+1 group, IgG concentrations were higher at day 28 than day 7 for serotypes 5, 7F, 9V, 14 18C and 23F (Supplementary Fig. 4B), while in the 1+1 group, IgG concentrations were similar between timepoints for most serotypes (except 6A and 6B; Fig. 2B).

For B_MEM,_ overall, there was a modest increase from baseline (12m) at day 7 for most serotypes with a further modest increase at day 28. B_MEM_ responses in both PCV10 schedules at day 7 and day 28 post-vaccination were higher than baseline for all serotypes except serotype 3 (at both timepoints) and for serotypes 19F and 4 (day 7 only) in the 0+1 group (Fig. 2A, Supplementary Fig. 4A). Comparing the 0+1 and 1+1 PCV10 groups, the 1+1 group had higher B_MEM_ responses for serotypes 5, 6B and 14 (Fig. 2A, Supplementary Fig. 4A) than the 0+1 group at day 7, while all other serotypes were similar. By day 28, B_MEM_ responses were similar for all serotypes between PCV10 schedules (Fig. 2A, Supplementary Fig. 4A). B_MEM_ for non-PCV10 serotypes (3, 6A, and 19A) were low and did not increase from baseline at day 7 or day 28 in both a 0+1 and 1+1 schedule (Supplementary Fig. 4A).

For PCV13 0+1, a modest increase in B_MEM_ was observed at day 7, similar to IgG, however unlike IgG, did not increase from day 7 at day 28. Similarly, for the PCV13 1+1 group, B_MEM_ increased from baseline at day 7, and remained similar at day 28. In terms of B_MEM_ comparing the 0+1 and 1+1 PCV13 groups, the 1+1 group had higher B_MEM_ responses for serotypes 18C and 23F (Supplementary Fig. 4B) than the 0+1 group at day 7, while all other serotypes were similar. By day 28, B_MEM_ responses were similar for all serotypes between PCV13 schedules (Fig. 2B; Supplementary Fig. 4B). Comparing day 7 to day 28 within each schedule, B_MEM_ responses in the 0+1 group were higher at day 28 for serotypes 3 and 23F, whereas in the 1+1 group only serotype 18C was higher (Supplementary Fig. 4B).

### Comparison of serotype-specific IgG and B_MEM_ between PCV10 and PCV13 schedules

Comparing the two 0+1 schedules, at day 7 post-vaccination the PCV13 0+1 schedule produced higher IgG concentrations than the PCV10 0+1 schedule for all serotypes, except 18C which was higher in the PCV10 0+1 group (Fig. 3A, Supplementary Fig. 5A). By day 28 post-vaccination, IgG responses were similar for most of the shared serotypes between PCV10 and PCV13 (except serotypes 4, 5 and 18C; Fig. 3A, Supplementary Fig. 5A).

**Figure 3.**
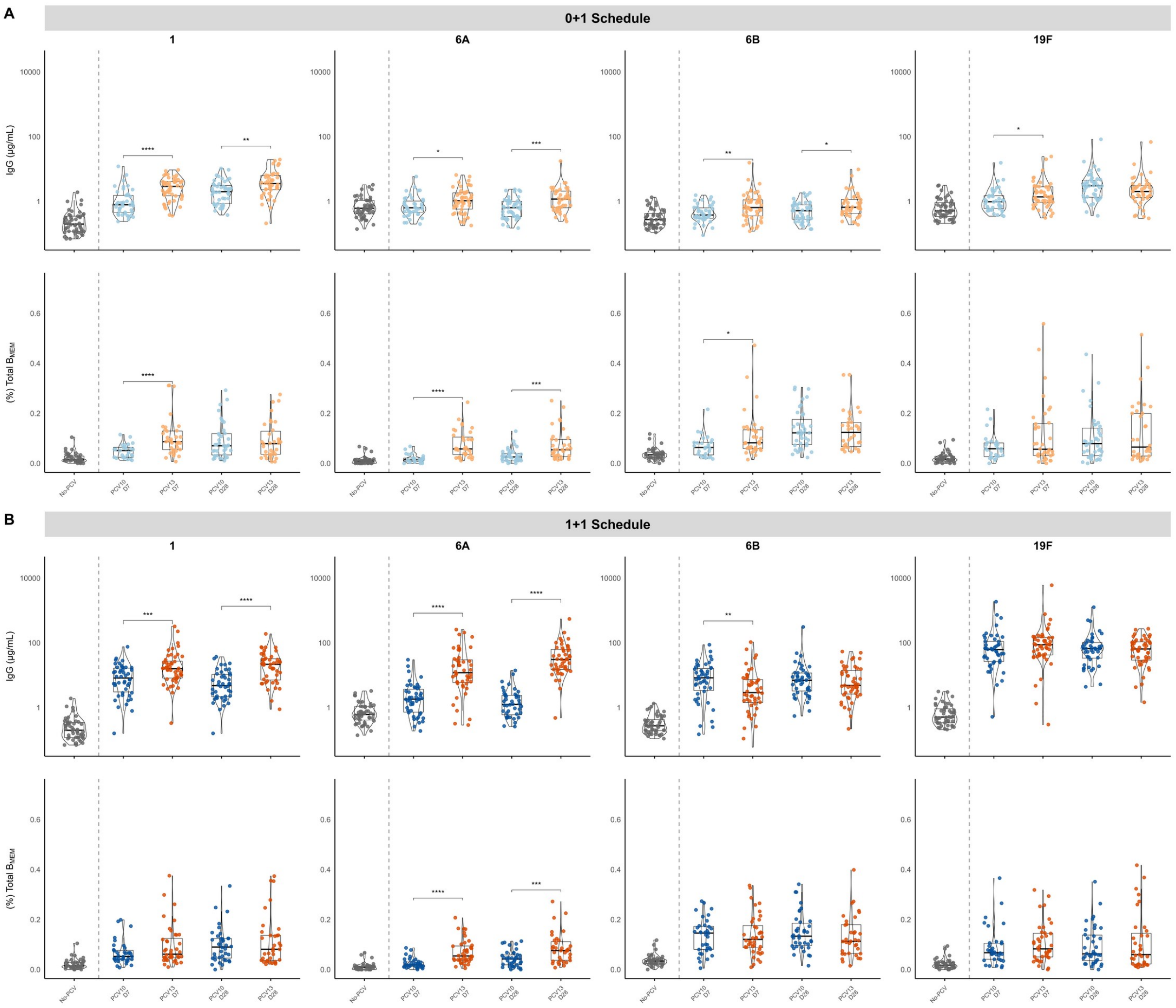
- Comparison of a 0+1 or 1+1 dosing schedule between PCV10 and PCV13 Serotype-specific B_MEM_ frequencies and serum IgG concentrations were measured at day 7 and day 28 post-12m following 0+1 and 1+1 dosing schedules. Data are shown for 0+1 (**A**) and 1+1 (**B**) dosing schedules. For both the PCV10 and PCV13 vaccinated groups, measurements are shown at day 7 and day 28 post-vaccination. A no-PCV group at 12m (grey) is shown. Serotype-specific B_MEM_ are expressed as a proportion of total B_MEM_, and serotype-specific IgG concentrations are expressed as µg/mL. Box plots indicate the median and interquartile range, with violins showing the distribution within each group. Statistical significance was assessed using the Mann–Whitney test and is displayed for comparisons between PCV10 and PCV13 at day 7 or day 28 post-vaccination. Significance is denoted as *p < 0.05, **p < 0.01, ***p < 0.005, ****p < 0.001.

Specific IgG for PCV13-only serotypes (3, 6A, and 19A) was higher in the PCV13 0+1 than PCV10 0+1 group at day 7 and day 28 (Supplementary Fig. 5A).

Similarly, the PCV13 0+1 group produced higher frequencies of B_MEM_ at day 7 for 7 of the 10 shared PCV10 serotypes (1, 4, 5, 6B, 7F, 14, and 18C; Fig. 3A, Supplementary Fig. 5A) as well as all PCV13-only serotypes (3, 6A, 19A), while serotypes 19F, 23F and 9V were similar, compared to PCV10. At day 28, both PCV13 and PCV10 0+1 schedules had similar B_MEM_ responses for almost all shared serotypes (except 18C; Supplementary Fig. 5A), while PCV13-only serotypes were higher in the PCV13 group (Fig. 3A, Supplementary Fig. 5A).

Comparing the two 1+1 schedules, at day 7 the PCV13 1+1 group produced higher IgG concentrations than the PCV10 1+1 group for 4/10 shared serotypes (1, 5, 7F, and 9V) and the PCV13-only serotypes 3, 6A, and 19A (Fig. 3, Supplementary Fig. 5B). The remaining serotypes had similar (4, 14, 19F, 18C, 23F; Fig. 3B, Supplementary Fig. 5B) or lower (6B; Fig. 3B) IgG concentrations. At day 28, IgG concentrations remained higher for 5/10 shared serotypes (1, 5, 7F, 9V and 23F; Fig. 3B; Supplementary Fig. 5B) and all PCV13-only serotypes in the PCV13 group, and were similar for 5 serotypes (6B, 19F, 4, 14, and 18C; Fig. 3B, Supplementary Fig. 5B).

Fewer differences were seen between vaccine products with B_MEM_ responses. B_MEM_ were higher in the 1+1 PCV13 group compared to 1+1 PCV10 group for shared serotypes 5 and 18C at day 7 (Supplementary Fig. 5B). B_MEM_ responses for PCV13-only serotypes were higher at day 7 in the PCV13 group (Fig. 3B, Supplementary Fig. 5B). At day 28, responses were similar for all shared serotypes except serotype 18C, which was higher for PCV13 (Supplementary Fig. 5B), and PCV13-only serotypes 3 (Supplementary Fig. 5B), and 6A but not 19A (Fig. 3B, Supplementary Fig. 5B).

### Magnitude of total serotype-specific B_MEM_ between 0+1 and 1+1 schedules of PCV10 and PCV13

Overall, both PCV10 and PCV13 produced robust B_MEM_ responses at day 7 and day 28 (Fig. 4A, Fig. 4B). Prior to the 12m dose, vaccine-type B_MEM_ represented 0.3-0.4% of total B_MEM_ cell population, but this increased to 1.2% of total B_MEM_ (Fig. 4A, B). For the 0+1 PCV10 group, the total B_MEM_ proportion (to all PCV10 serotypes plus non-vaccine serotypes 3, 6A and 19A) increased ∼2-fold by day 7 and ∼3-fold by day 28 compared to baseline (this was a ∼2-fold and 3.5-fold increase for PCV10 only serotypes; Fig. 4A). For the 1+1 PCV10 group, the total B_MEM_ proportion increased ∼2-fold by day 7 and 2.4-fold by day 28 compared to baseline (∼2.3-fold by day 7 and ∼2.7-fold by day 28 for PCV10-only serotypes; Fig. 4A).

**Figure 4.**
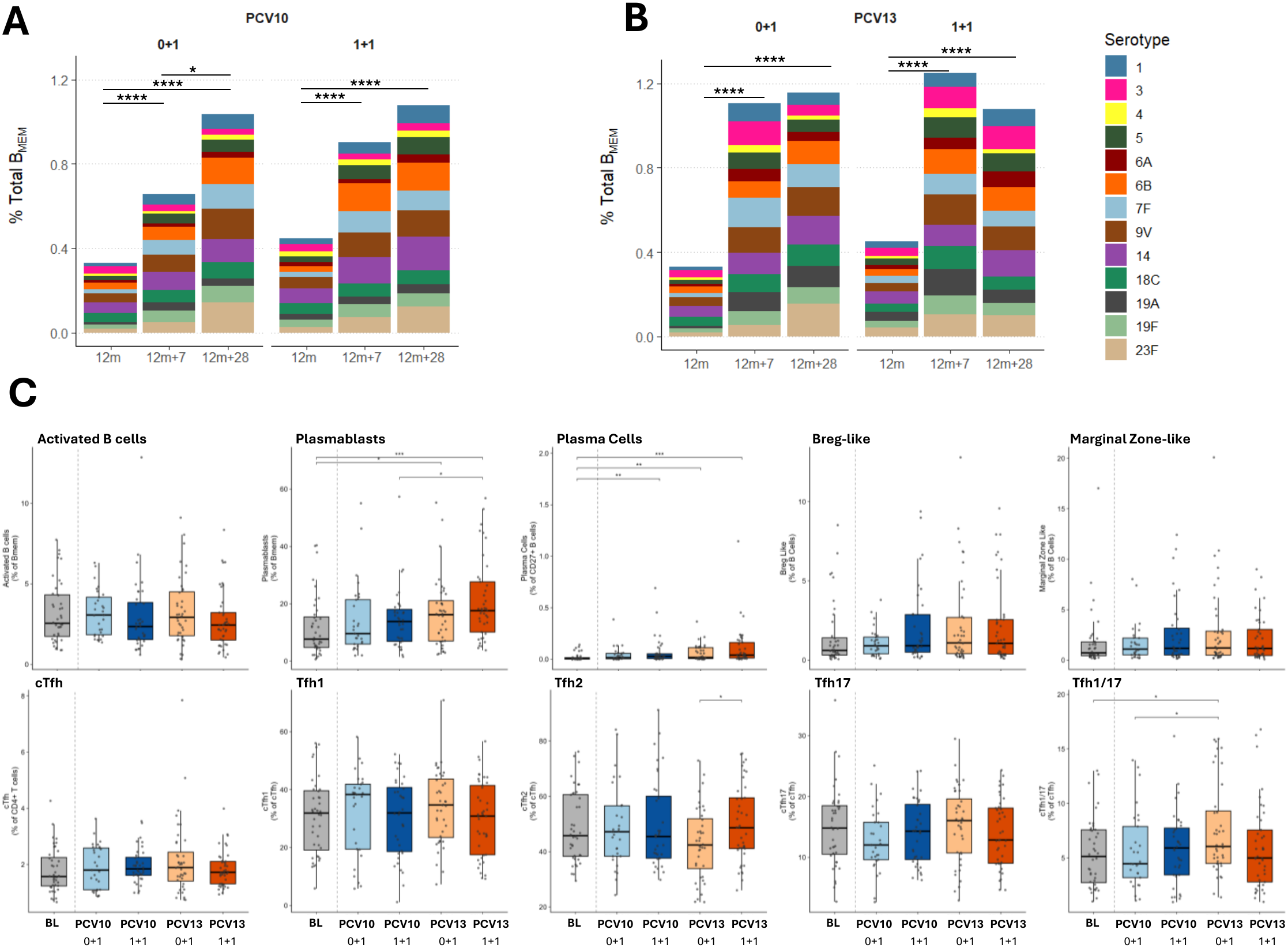
- Cellular immune changes following 0+1 and 1+1 schedules of PCV10 or PCV13. Cumulative serotype-specific B_MEM_ responses following PCV10 (**A**) or PCV13 (**B**) vaccination in Vietnamese infants. Measurement of B cell populations including activated B cells, plasmablasts, plasma cells Breg-like cells and marginal zone-like cells and T cell populations including circulating T-follicular helper cells (cTfh) as well as Tfh1, Tfh2, Tfh17, Tfh1/17 populations (**C**) were also undertaken. Activated B cells were calculated as a proportion of total circulating B cells, plasmablasts as a proportion of B_MEM_ and plasma cells as a proportion of CD27^+^ B cells. For T cells, cTfh were calculated as a proportion of CD4^+^T cells, and Tfh1, Tfh2, Tfh17, Tfh1/17 as proportions of total cTfh. Baseline at 12m (grey), PCV10 0+1 (light blue), PCV10 1+1 (dark blue), PCV13 0+1 (light orange) and PCV13 1+1 (dark orange) groups from 7-days post-vaccination. Significance is denoted as *p < 0.05, **p < 0.01, ***p < 0.005, ****p < 0.001 from unpaired Wilcoxon tests.

For PCV13, there was a ∼3.2-fold increase at day 7 and ∼3.5-fold increase at day 28 in B_MEM_ from baseline. For the PCV13 1+1 schedule, B_MEM_ responses peaked at day 7 (∼2.6-fold) and were similar by day 28 (∼2.4-fold) post-vaccination (Fig. 4B).

### Memory B cell and cTfh responses following PCV vaccination

As peak cellular responses tend to occur by 1-week post-vaccination, we examined major memory cell populations induced post-PCV vaccination.

For PCV10 at day 7, there were no differences between activated Bmem between any of the study groups. Compared to baseline, plasmablasts were highest in the PCV13 1+1 group at day 7, followed by the PCV13 0+1 group. Similarly, for PCV10, plasmablasts were elevated from baseline but not different for either PCV group but were lower compared to PCV13.

Plasma cells were also highest in the PCV13 1+1 group and PCV10 0+1 group compared to baseline but no differences between either vaccine or schedule was observed. Breg-like cells were elevated in the PCV10 0+1 group compared to baseline and were higher than the PCV10 1+1 group. No differences were found for both PCV13 groups, although median levels were above baseline. MZ-like B cells were lower in both PCV10 groups with greatest reduction for the 1+1 group compared to baseline. No differences were seen for both PCV13 groups compared to baseline, although PCV13 1+1 group had higher MZ B cells than PCV10 1+1 group (Fig. 4C). At day 28, activated B_MEM_ were highest in a PCV13 0+1 group. Similarly, Breg-like cells were highest in PCV13 0+1 group compared to PCV10 0+1. Plasma cells, plasmablasts and MZ-like B cells were similar across groups and not different compared to baseline (Supplementary Fig. 6).

For total cTfh, there were no differences between either PCV or schedule at day 7. Tfh1 and Tfh17 were also similar between groups and were not different from baseline. For PCV13, Tfh2 was lower in the 0+1 compared to 1+1 schedule, but similar to baseline. Tfh1/17 cells were higher in PCV13 0+1 than baseline and compared to the PCV10 0+1 group (Fig. 4C). At day 28, cTfh levels were higher than baseline for PCV13 1+1. Tfh17 cells were lower than baseline in PCV13 0+1 but similar for to other groups. Tfh1, Tfh2 and Tfh1/17 subsets were similar across groups and compared to baseline (Supplementary Fig. 6).

### Serotype-specific Bmem in seroprotected compared with non-seroprotected infants

We found a substantial proportion of infants did not have seroprotective IgG levels (≥0.35 μg/mL) at day 7 or day 28 post-vaccination for serotypes 6B (>75%) and 23F (>50%). This was mostly in the 0+1 PCV10 or 0+1 PCV13 groups (Supplementary Fig. 7). For these infants, we wanted to determine whether B_MEM_ for those serotypes could be generated by a 0+1 schedule (Fig. 5). We found that B_MEM_ responses for serotypes 6B and 23F were similar between infants who did or did not have seroprotective IgG levels at day 7 and day 28. The exception was serotype 23F where B_MEM_ responses at day 7 were lower among infants who had IgG less than 0.35 μg/mL, but this was similar by day 28 (Fig. 5).

**Figure 5.**
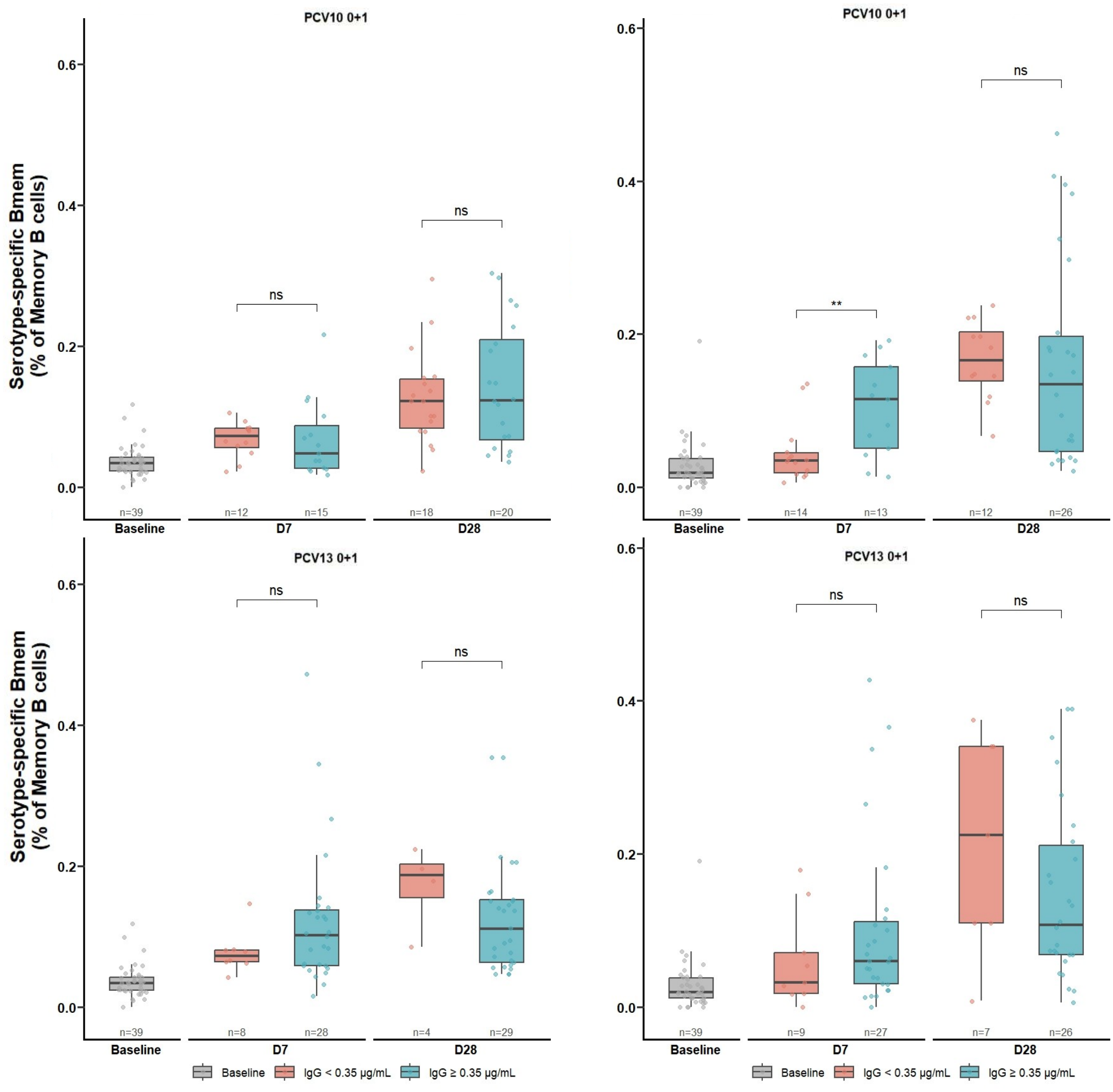
- Comparison of serotype-specific B_MEM_ responses among children who do or do not generate seroprotective antibody responses ≥ 0.35µg/mL Serotype-specific B_MEM_ in participants in the PCV10 and PCV13 0+1 groups at baseline (grey), or day 7 post-vaccination (top) or day 28 post-vaccination (bottom) with IgG < 0.35 µg/mL (red) and ≥ 0.35 µg/mL (blue) for serotypes 6B (left) and 23F (right). Ns = not significant, **p < 0.01.

### Antibody isotype and subclass distribution of serotype-specific B_MEM_

The antibody isotype and subclass profiles of these serotype-specific B_MEM_ populations at day 7 post-vaccination (Fig. 6) were also determined for those serotypes where sufficient B_MEM_ numbers were available. Isotype and subclass profiles were not determined at day 28, as B_MEM_ responses were mostly similar. Overall, the IgG, IgM and IgA isotype distribution was similar between the two PCVs and schedules, with some exceptions. The PCV10 groups produced more unswitched (IgD^+^IgM^+^) serotype-specific B_MEM_ for serotype 1, whereas for serotype 23F, the 1+1 schedule produced more IgM^+^ B_MEM_ than the 0+1 which produced higher proportions of unswitched B_MEM_ (Fig. 6A, Supplementary Fig. 7). The PCV13 1+1 schedule generated a higher proportion of switched serotype-specific B_MEM_ (correspondingly fewer unswitched, IgD^+^IgM^+/-^ cells) than the PCV13 0+1 schedule (Fig. 6A, Supplementary Fig. 7). The PCV13 1+1 schedule produced more IgG^+^B_MEM_ for serotypes 1, 5, 9V, 18C, 19A and 19F compared to PCV10 1+1, whereas PCV13 0+1 and PCV10 0+1 groups showed similar levels of IgG^+^B_MEM_. Most of these serotype-specific IgG^+^ B_MEM_ were of the IgG1 subclass, accounting for >70% of the IgG^+^B_MEM_ across both PCVs and schedules. Minimal proportions for IgG2^+^ B_MEM_ and IgG3^+^B_MEM_ were observed, except for the non-vaccine type 19A (∼50%) and 23F (∼30%) in the 1+1 PCV10 schedule for IgG3^+^B_MEM_ (Fig. 6B).

**Figure 6.**
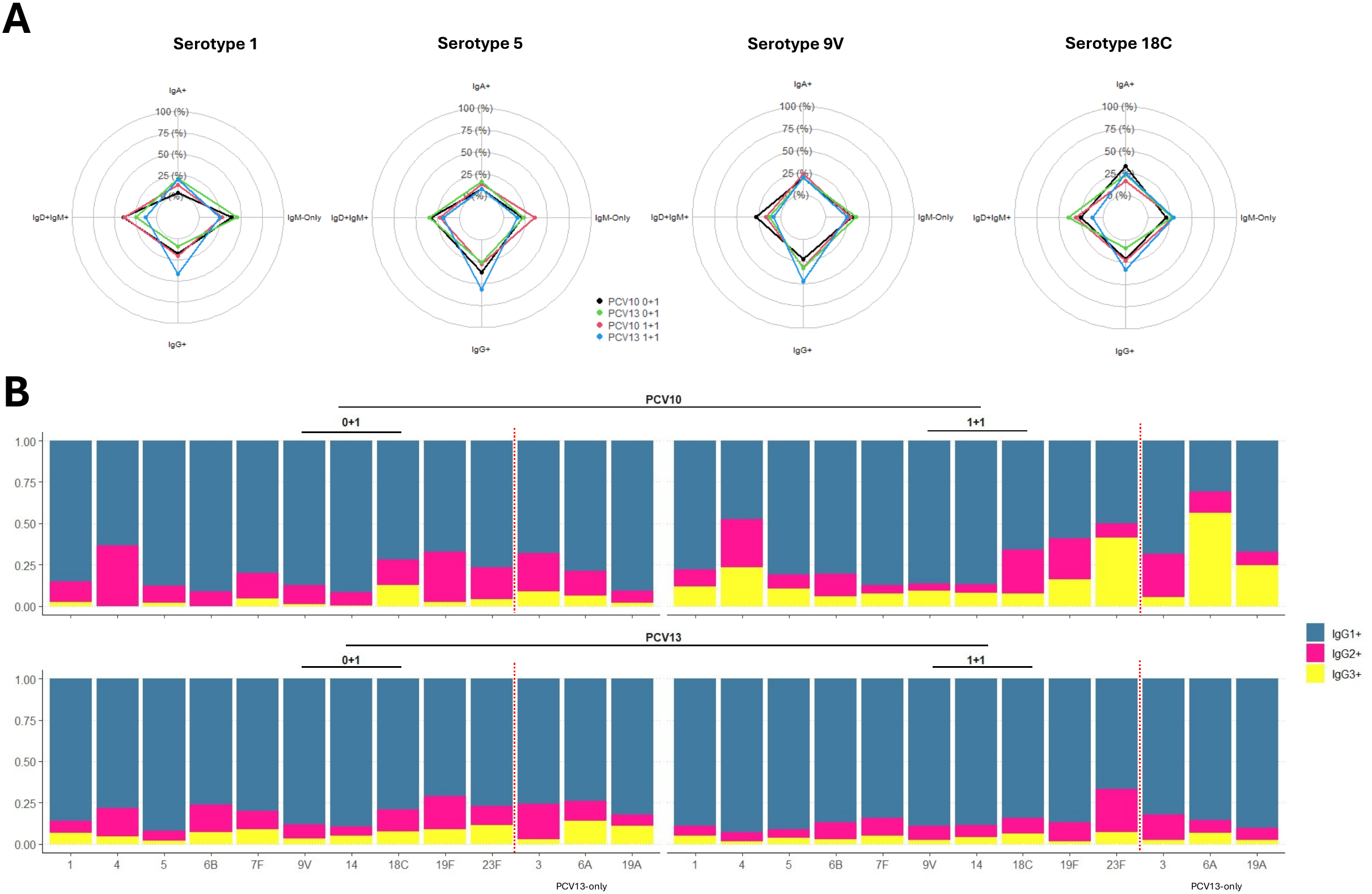
- Serotype-specific isotype and subclass distribution **A**, Median isotype distribution for serotype-specific B_MEM_ populations for representative serotypes 1 (top left), 5 (top right), 9V (bottom left), and 18C (bottom right). **B**, Stacked bar graph of IgG subclass distribution by serotype.

## Discussion

Reduced-dose (1+1) PCV schedules are now endorsed by the WHO for children under 5 years of age in settings with a well-established PCV programme (>5 years), high coverage (≥80%) and low vaccine-serotype circulation ^5^. These schedules rely on herd protection (carriage) through reduced nasopharyngeal carriage. Memory B cell responses are thought to provide long-term protection by PCVs, but no data exists from 1+1 RCTs in infants. For this analysis, we used spectral flow cytometry to comprehensively measure serotype-specific B_MEM_ responses to all vaccine-serotypes following 1+1 and 0+1 schedules of PCV10 and PCV13 in Vietnamese infants. Both PCVs and schedules induced robust B_MEM_ responses, with PCV13 generating higher B_MEM_ than PCV10. Stronger B_MEM_ responses were also induced by the 1+1 schedule for both PCVs by day 7 but were similar by day 28 compared to the 0+1 schedule. Furthermore, PCV13 B_MEM_ responses were associated with increased plasmablasts at day 7 and cTfh by day 28 but not for PCV10. Our findings suggest that PCV10 and PCV13 differ in the kinetics and magnitude of B_MEM_ induction, which may have implications for long-term protection.

Serotype-specific B_MEM_ have been associated with protection against carriage in adult human challenge studies ^18^. However, data from infant 1+1 schedules are lacking ^9^. Our finding of stronger Bmem for PCV13 is consistent with previous post-booster studies using more than 2 doses ^19^. Differences in B_MEM_ between PCV13 and PCV10 may reflect variation in carrier protein, conjugation chemistry or antigen composition, all of which are known to influence B cell activation and memory formation ^20,21^. For both PCVs, enhanced B_MEM_ in the 1+1 schedule at day 7 likely reflects the presence of primed antigen-specific B_MEM_, enabling accelerated expansion upon boosting. In contrast, the 0+1 PCV10 schedule generated a delayed B_MEM_ response compared to PCV13 but was comparable to the 1+1 schedule by day 28. This is important for the use of PCV in catch-up campaigns or in emergency settings ^22^, where only 1 dose of PCV might be given.

The increased PCV13 B_MEM_ responses in the 1+1 group were also associated with higher plasmablast and plasma cell numbers by day 7, as well as higher cTfh by day 28, supporting the generally higher immunogenicity observed with PCV13 in this study and others ^23^. This could suggest different germinal centre engagement by these two vaccines ^24,25^. In our parent trial, PCV13 had a stronger impact on carriage compared to PCV10 in a 1+1 schedule, consistent with our B_MEM_ data. However, a 0+1 PCV10 schedule, which produced lower IgG but similar B_MEM_ to PCV13, had a greater impact on carriage by 24 months of age (53% v 32%) ^11^. This suggests that the relative contributions of serotype-specific IgG and B_MEM_ to protection might differ based on the schedule the vaccine is given. Despite both PCVs being highly protective, our data provide additional insights into how they mediate such protection.

The role of pneumococcal B_MEM_ in PCV efficacy is supported by trial data showing that protection against carriage occurs even after antibodies have waned ^7^. B_MEM_ are rarely studied as part of infant PCV RCTs ^27^. In adults, PCVs induce robust B_MEM_ responses following multiple doses ^19,28–30^. The divergent kinetics of IgG and B_MEM_ that we observed are consistent with other vaccines, in which B_MEM_ frequencies are maintained independently of antibody titres ^31–33^. An interesting finding from our study was that B_MEM_ for serotypes 6B and 23F were detected even among infants who had IgG levels below the seroprotective correlate of protection (0.35 µg/mL), suggesting that these infants are likely to be protected over the long-term. An important caveat of this result is that the IgG threshold is an aggregate population-level correlate and is less reliable at the individual-level ^34^.

We used spectral flow cytometry to allow direct *in vivo* enumeration of serotype-specific B_MEM_ for all vaccine serotypes, providing unprecedented immunological insights following reduced dose PCV schedules in infants. Previous studies in adults and infants have mostly relied on *in vitro* expansion of B cells in culture by ELISpot, including our previous study from this trial. While ELISpot is considered a gold-standard method ^35^, it may selectively promote the differentiation of certain subsets of B_MEM_ and therefore not capture the full spectrum of B_MEM_ responses following PCV ^9^. Our method also allowed phenotyping of other B cell and Tfh populations important in memory formation following vaccination that have been measured in adult PCV studies but not infant 1+1 studies ^9^.

The relationship between PCV immunity and protection against carriage in children has not been established. Serotype-specific IgG thresholds for carriage are typically higher than the 0.35 µg/mL threshold used for IPD and differ between serotypes and settings ^34,36,37^. Our earlier PCV trial in Vietnam found no association between serotype-specific B_MEM_, IgG or OPA at 10 months of age following a 3+0, 3+1 or 2+1 PCV10 schedule and carriage at 18 or 24 months of age ^28^. Nevertheless, the relationship between IgG and B_MEM_ with carriage outcomes remains an open question and becomes particularly important as countries consider using a 1+1 schedule. For B_MEM_, the only evidence to date comes from human challenge studies for serotype 6B ^16^. While informative, this is likely to be very different to what occurs in children, especially those living in high-burden settings. Analyses from this RCT to examine the relationship between IgG and B_MEM_ and long-term carriage acquisition is ongoing and will provide further insights into these mechanisms ^11^.

This RCT was conducted in a setting with substantial pneumococcal burden and no infant PCV programme, allowing comprehensive assessment of PCV effects, showing that both the 0+1 and 1+1 schedules of PCV10 and PCV13 had a major impact of vaccine-type carriage (up to 65%) compared to unvaccinated infants by 24 months of age. A key strength of our study was the analysis of B_MEM_ and other immune cell populations in infants under RCT settings. This provides the strongest data to date for unravelling PCV immunogenicity and memory following a 0+1 or 1+1 schedule. We also assessed responses at day 7 and day 28 to gain a full understanding of the PCV response in the month following vaccination. Our study also had some important limitations. First, our B_MEM_ data was from different sets of randomised infants at each timepoint, which limited our ability to undertake pairwise analyses and may have underestimated some of the effects observed. The relationship with B_MEM_ and carriage protection is ongoing and will be an important next step to understand correlates of protection. Finally, while our results suggest robust B_MEM_ by PCV10 and PCV13, this cannot be generalised to other vaccines such as Pneumosil PCV10 or higher-valency products ^38^.

In conclusion, this study demonstrates that reduced-dose PCV schedules induce robust serotype-specific B_MEM_ responses in infants. Together, these data support B_MEM_ as an important measure of vaccine-induced immunity in addition to serology. Establishing the relationship between B_MEM_ responses and protection against carriage in the second year of life will be a critical next step in determining correlates of protection and informing pneumococcal vaccine policy.

## Data Availability

All data produced in the present work are contained in the manuscript.

## Acknowledgments

We thank the study participants and their families, and all VPT-II study and laboratory staff from the Pasteur Institute of Ho Chi Minh City, the Murdoch Children’s Research Institute, and the Menzies School of Health Research. We acknowledge the contributions of Keith Klugman and Gail Rodgers. This work was supported by the Bill & Melinda Gates Foundation (grant number INV-008627) and a NHMRC CRE for Pneumococcal Control in the Asia-Pacific grant (GNT1196415). The funders contributed to refinement of the study design but had no role in data collection, data analysis, data interpretation, or writing of the report. The Murdoch Children’s Research Institute is supported by the Victorian Government’s Operational Infrastructure Support Program (applicable for all Murdoch Children’s Research Institute authors).

## Author contributions

PVL conceived this study of PCV B_MEM_ responses within the larger RCT in Vietnam. HEB conducted the B_MEM_ assays, analysed the data and wrote the first draft of the manuscript. VTTD undertook the IgG assays in Viet Nam, with oversight from RAH and PVL. HPT, BT and KB were involved in the establishment, day-to-day management, and implementation of the trial. CDN advised on the sample size and statistical analysis. TVN was involved in the design and establishment and had overall responsibility for the conduct of the trial in Viet Nam as the site principal investigator. KM conceived the overall VPT-II trial and had overall responsibility of the trial as the principal investigator. RAH, LQ, SL, and NM performed serological assays and assisted with the flow cytometry. JA provided supervision and oversight of the flow cytometry work. All authors contributed to refinement of and approved the submitted manuscript.

