## Supplementary Figures and Tables for "PNEUMOCOCCAL CONJUGATE VACCINE SCHEDULES SHAPE SEROTYPE-SPECIFIC MEMORY B-CELL RESPONSES"

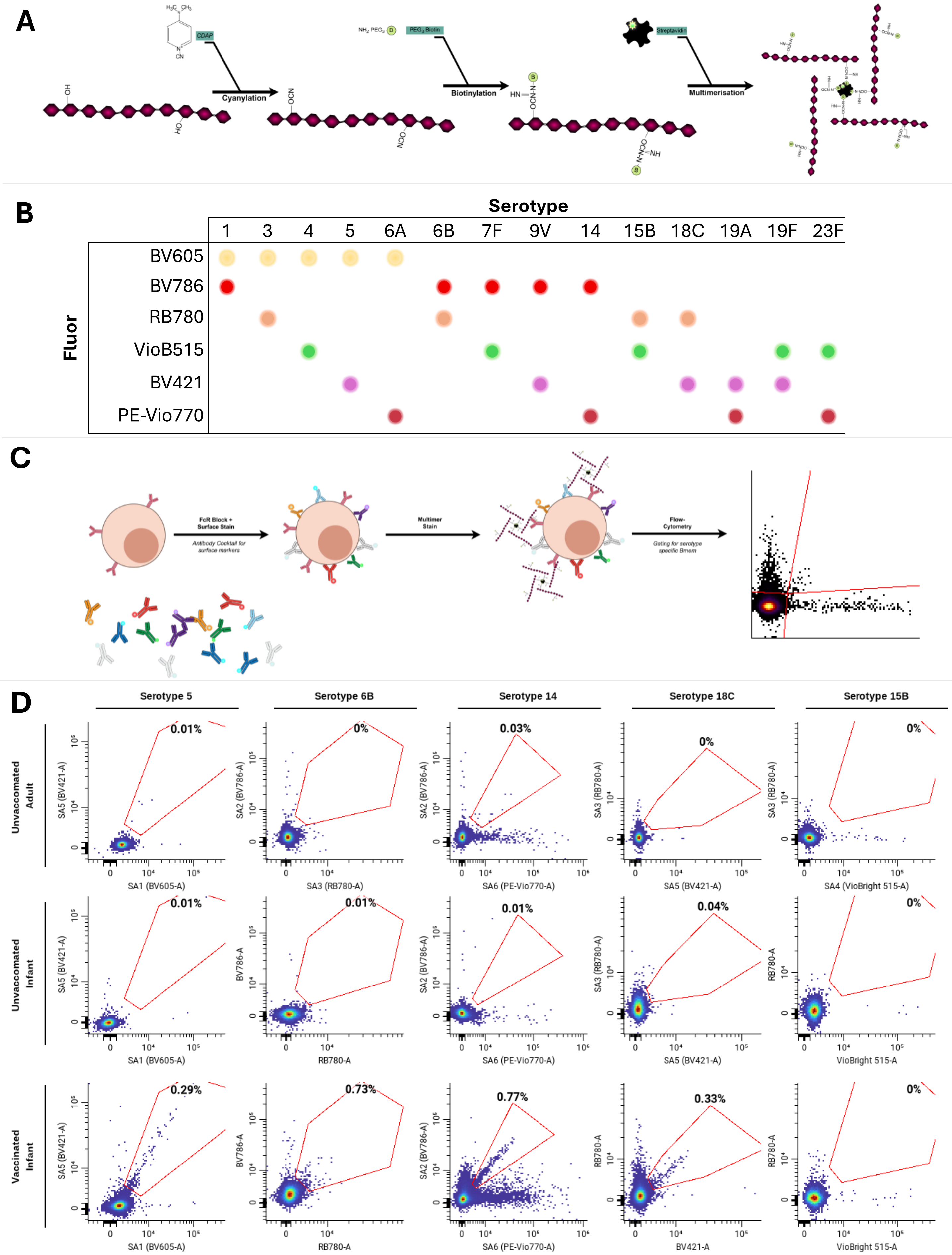


**S. Figure 1 - Study Workflow**

**A**, generalised workflow for modifying purified pneumococcal polysaccharides for biotinylation, and subsequent multimer formation. CDAP: 1-cyano-4-dimethylaminopryridine tetrafluoroborate. **B**, barcoding strategy showing fluorophore combinations for the streptavidin-polysaccharide multimers. **C**, basic staining workflow using B-cell immunophenotyping panel and multimers (left), and serotype-specific gating highlighting double positive selection (right).

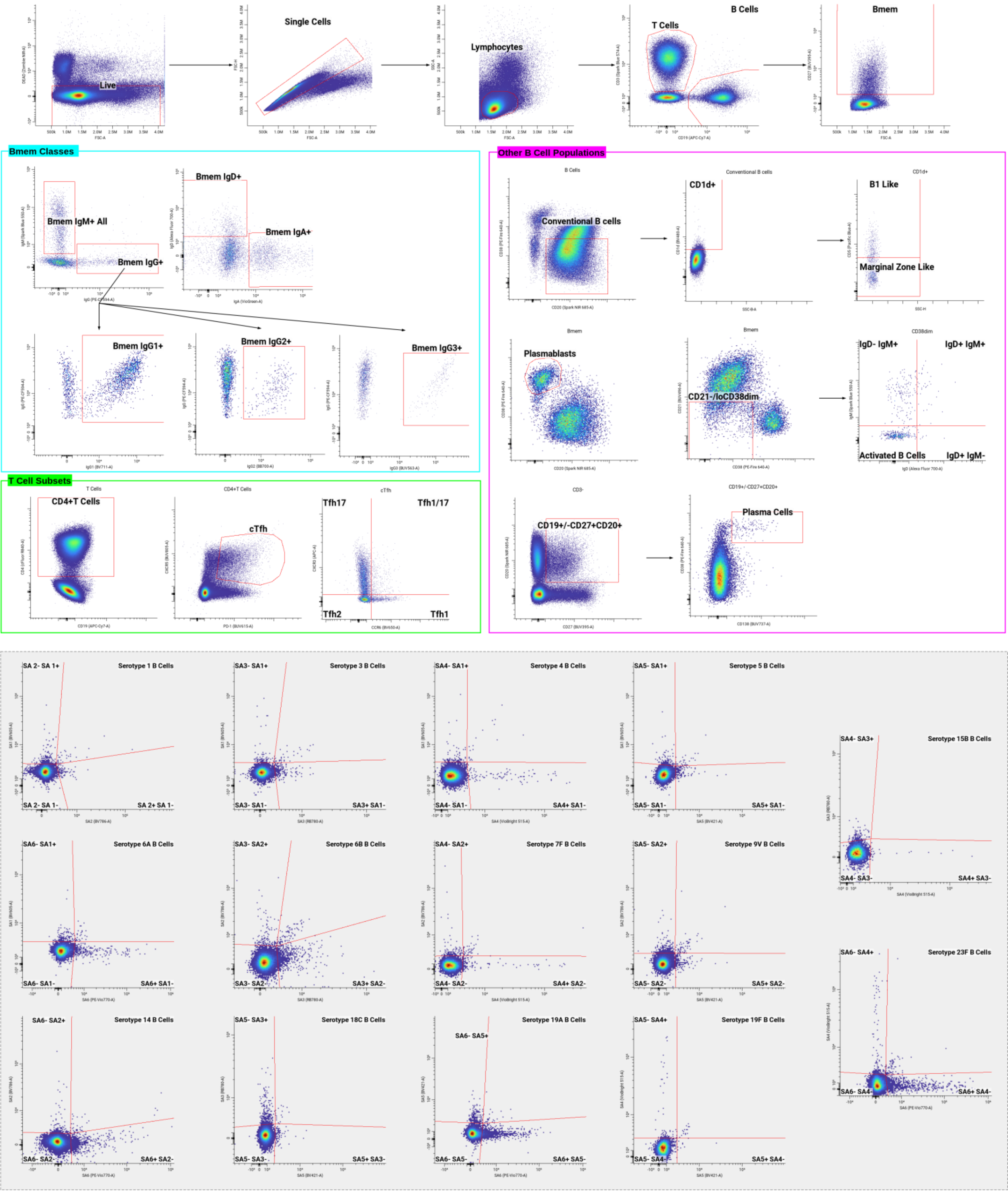


**S. Figure 2 – Gating Strategy**

Manual gating strategy to identify $IgG^+B_{MEM}$ ($CD3^-CD19^+CD27^+IgG^+$), $IgM^+B_{MEM}$ ($CD3^-CD19^+CD27^+IgM^+$), $IgD^+B_{MEM}$ ($CD3^-CD19^+CD27^+IgD^+$), $IgA^+B_{MEM}$ ($CD3^-CD19^+CD27^+IgA^+$), B1-like B cells ($CD3^-CD19^+CD38^{-/lo}CD20^+CD1d^+CD5^+$), marginal zone-like B cells ($CD3^-CD19^+CD38^{-/lo}CD20^+CD1d^+CD5^-$), plasmablasts ($CD3^-CD19^+CD27^+CD38^{hi}CD20^-$), plasma cells ($CD3^-CD20^+CD27^+CD38^{hi}CD138^+$), circulating Tfh ($CD3^+CD19^-CD4^+CXCR5^+PD\text{-}1^+$) and its subpopulations Tfh1 ($CCR6^+CXCR3^-$), Tfh2 ($CCR6^-CXCR3^-$), Tfh17 ($CCR6^-CXCR3^+$), Tfh1/17 ($CCR6^+CXCR3^+$), and serotype-specific $B_{MEM}$.

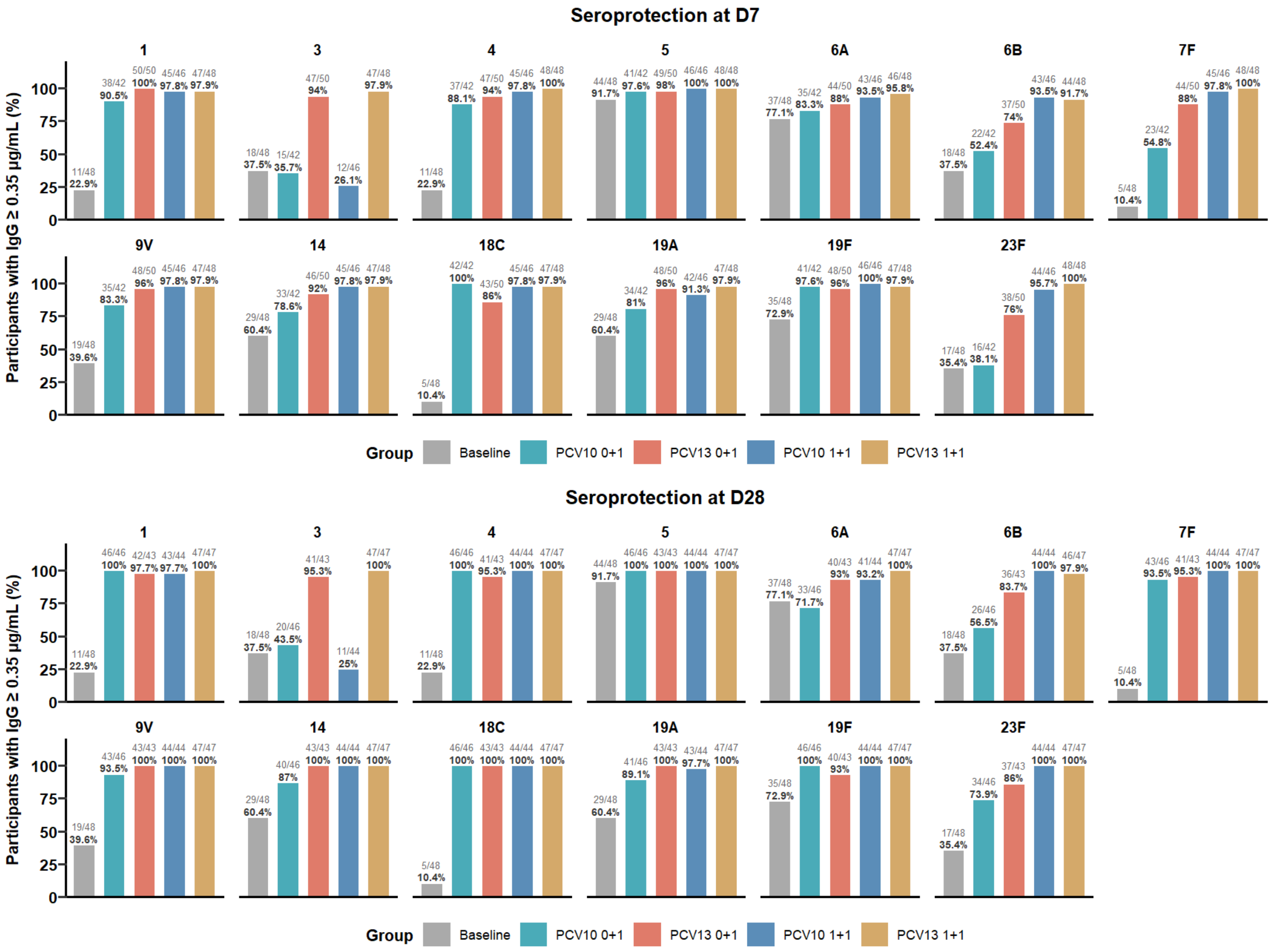


**S. Figure 3 – IgG sero-protection by serotype and vaccine group**

Bars display proportion of individuals (n/N; %) with IgG concentrations ≥ 0.35 µg/mL for each serotype at day 7 (top) and day 28 (bottom) post-vaccination. Baseline (grey) unvaccinated group at 12-months, PCV10 0+1 (turquoise), PCV13 0+1 (orange), PCV10 1+1 (blue), and PCV13 1+1 (yellow).

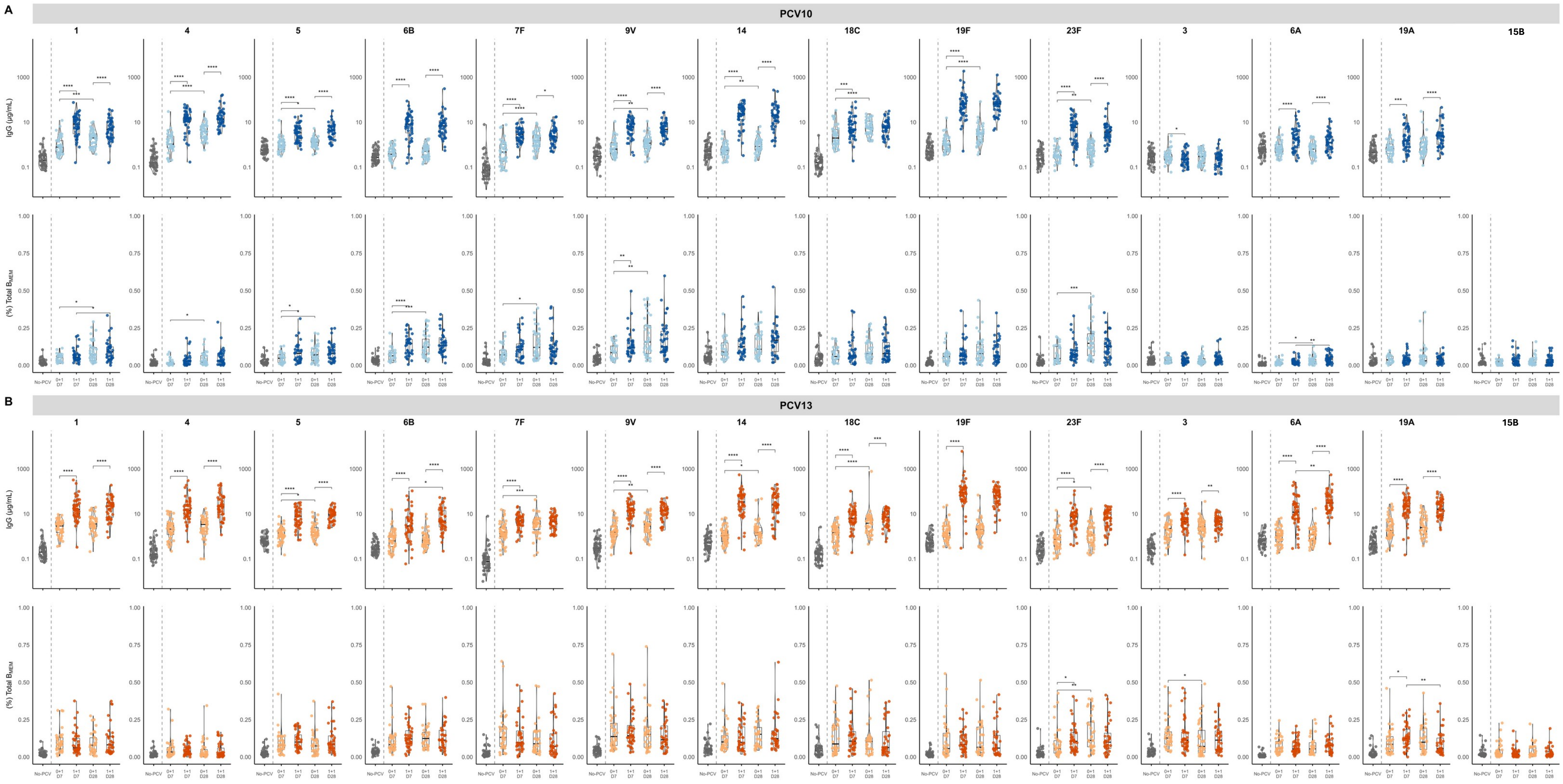


**S. Figure 4 - Comparison of 1+1 and 0+1 dosing schedules with (A) PCV10 and (B) PCV13**

Serotype-specific $B_{MEM}$ frequencies and serum IgG concentrations were measured at 7- and 28-days post-12m following 0+1 and 1+1 dosing schedules. Data are shown for PCV10 (**A**) and PCV13 (**B**). For the 0+1 schedule, measurements are shown at day 7 (yellow) and day 28 (blue); for the 1+1 schedule, measurements are shown at day 7 (green) and day 28 (pink). A no-PCV group at 12-months (red) is shown. Serotype-specific $B_{MEM}$ are expressed as a proportion of total $B_{MEM}$, and serotype-specific IgG concentrations are expressed as µg/mL in serum. Box plots indicate the median and interquartile range, with violins showing the distribution within each group. Statistical significance was assessed using the Mann–Whitney test and is displayed for comparisons between 0+1 and 1+1 schedules at day 7 or day 28 post-boost, and between day 7 and day 28 post-vaccination within each dosing schedule. Significance is denoted as *p < 0.05, **p < 0.01, ***p < 0.005, ****p < 0.001.

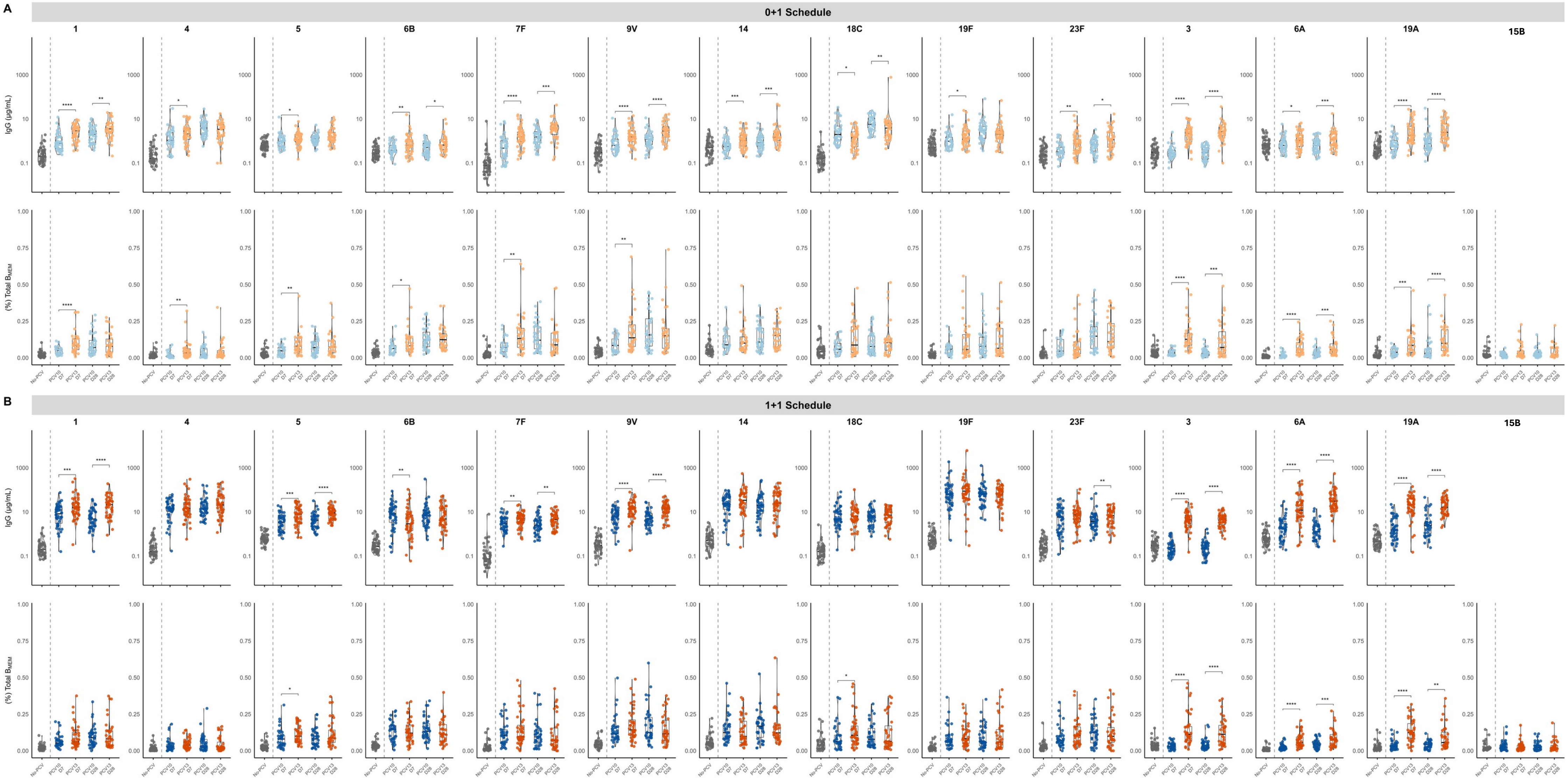


**S. Figure 5 - Comparison of a 0+1 or 1+1 dosing schedule between PCV10 and PCV13**

Serotype-specific BMEM frequencies and serum IgG concentrations were measured at day 7 and day 28 post-12m following 0+1 and 1+1 dosing schedules. Data are shown for 0+1 (A) and 1+1 (B) dosing schedules. For the PCV10 vaccinated, measurements are shown at day 7 (yellow) and day 28 (blue); for the PCV13 vaccinated, measurements are shown at day 7 (green) and day 28 (pink). A no-PCV group at 12m (red) is shown. Serotype-specific BMEM are expressed as a proportion of total BMEM, and serotype-specific IgG concentrations are expressed as μg/mL in serum. Box plots indicate the median and interquartile range, with violins showing the distribution within each group. Statistical significance was assessed using the Mann–Whitney test and is displayed for comparisons between PCV10 and PCV13 at day 7 or day 28 post-boost. Significance is denoted as *p < 0.05, **p < 0.01, ***p < 0.005, ****p < 0.001.

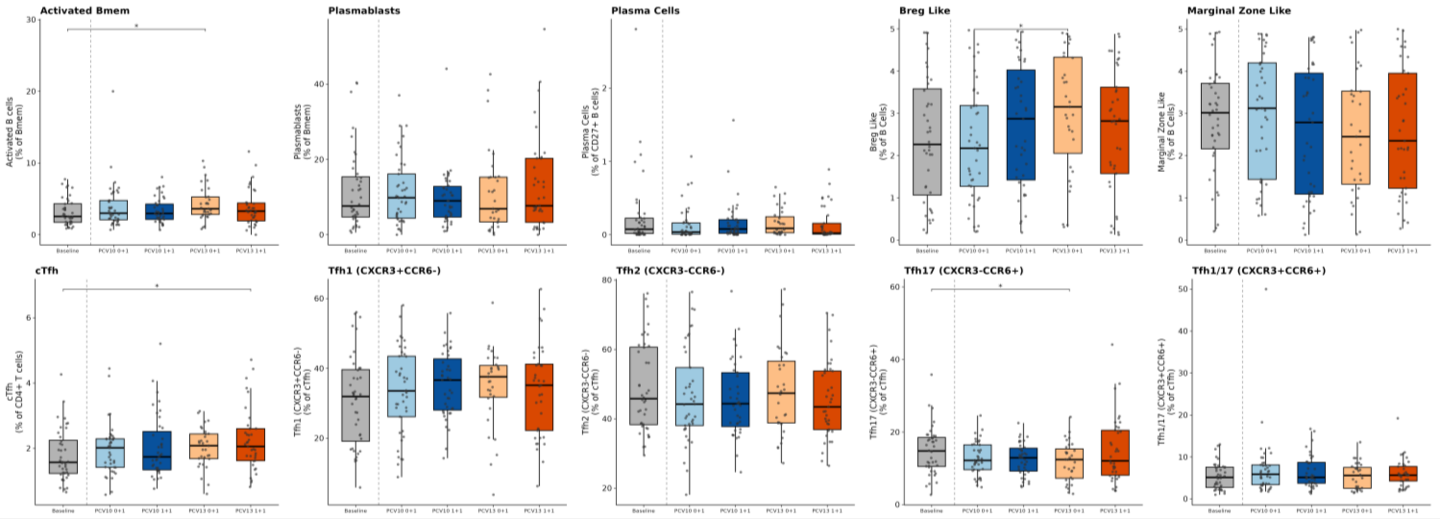


**S. Figure 6 – B cell and Tfh Populations 28-days post vaccination**

Activated B cells, Breg-like cells, and marginal zone-like cells are a proportion of total circulating B cells, plasmablasts as a proportion of $B_{MEM}$, plasma cells as a proportion of $CD27^+$ B cells, circulating T-follicular helper cells (cTfh) as a proportion of $CD4^+$T cells, and Tfh1, Tfh2, Tfh17, Tfh1/17 as proportions of total cTfh. Baseline at 12m (grey), PCV10 0+1 (light blue), PCV10 1+1 (dark blue), PCV13 0+1 (light orange) and PCV13 1+1 (dark orange) groups from 28-days post-vaccination. Significance is denoted as *p < 0.05, **p < 0.01, ***p < 0.005, ****p < 0.001 from unpaired Wilcox tests.

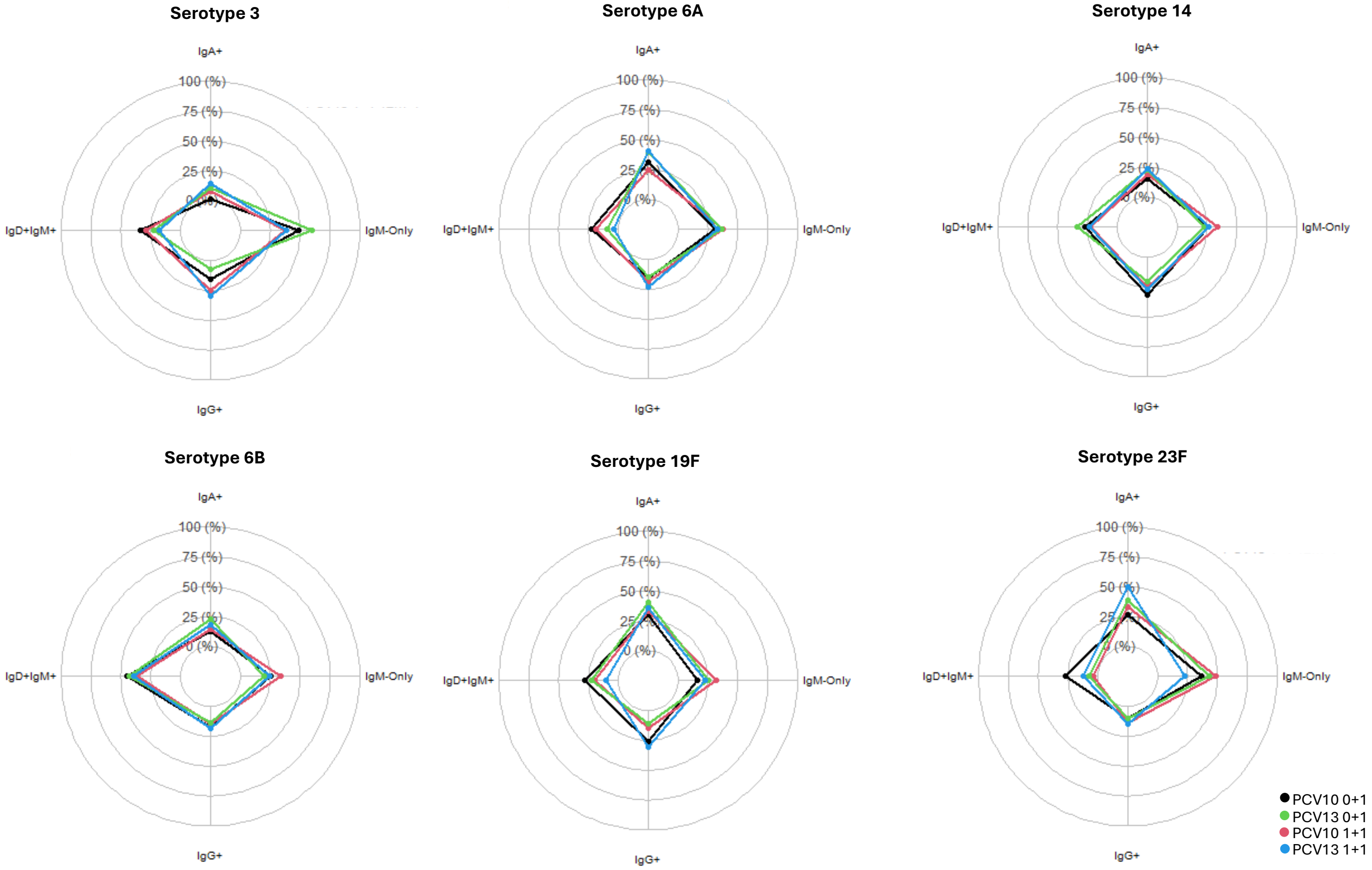


**S. Figure 7 – Serotype-specific $B_{MEM}$ Isotype distribution**

Median isotype distribution for serotype-specific $B_{MEM}$ populations for PCV serotypes at day 7 post-vaccination. PCV10 0+1 (black), PCV10 1+1 (red), PCV13 0+1 (green) and PCV13 1+1 (blue).

*Supplementary Table 1*

| Product | Supplier | Cat Number |
|---|---|---|
| **CWPS** | SSI Diagnostica | 68866 |
| **Amine-PEG3-Biotin** | Thermo Fisher Scientific | 21347 |
| **DMAP** | Sigma Aldrich | 8510550025 |
| **CDAP** | Merck (Sigma Aldrich) | SBR00023-1ML |
| **D-biotin** | Merck (Sigma Aldrich) | B4501 |
| **Foetal Bovine Serum (FBS)** | MCRI Internal Supply | |
| **Anti-pneumococcal capsule Reference Serum** | NIBSC | 007sp |
| **100 kDa Amicon ultracentrifugation unit** | Merck (Sigma Aldrich) | UFC510024 |
| **Ficoll Paque Plus** | Merck (Sigma Aldrich) | GE17-1440-02 |
| **Goat anti-human IgG** | Sigma Aldrich | A9544 |
| **Serotype 1 purified polysaccharide (PPS)** | ATCC | 161-X |
| **Serotype 3 PPS** | ATCC | 32-X |
| **Serotype 4 PPS** | ATCC | 35-X |
| **Serotype 5 PPS** | ATCC | 38-X |
| **Serotype 6A PPS** | ATCC | 271-X |
| **Serotype 6B PPS** | ATCC | 107-X |
| **Serotype 7F PPS** | ATCC | 281-X |
| **Serotype 9V PPS** | ATCC | 325-X |
| **Serotype 14 PPS** | ATCC | 74-X |
| **Serotype 15B PPS** | ATCC | 519-X |
| **Serotype 18C PPS** | ATCC | 289-X |
| **Serotype 19A PPS** | ATCC | 249-X |
| **Serotype 19F PPS** | ATCC | 99-X |
| **Serotype 22F PPS** | ATCC | 529-X |
| **Serotype 23F PPS** | ATCC | 217-X |
| **Serotype 33F PPS** | ATCC | 534-X |

*Supplementary Table 2*

| Marker | Fluorochrome | Manufacturer | Catalogue Number | Stain |
|---|---|---|---|---|
| **CXCR3** | APC | BD Biosciences | 561732 | 1 |
| **PD1** | BUV615 | BD Biosciences | 612991 | 1 |
| **CCR6** | BV650 | BD Biosciences | 563922 | 1 |
| **CD27** | BUV395 | BD Biosciences | 563815 | 1 |
| **CXCR5** | BUV805 | BD Biosciences | 741980 | 1 |
| **IgM** | Spark Blue 550 | BioLegend | 314556 | 2 |
| **CD3** | Spark Blue574 | BioLegend | 300488 | 2 |
| **IgG2** | BB700 | BD Biosciences | 756059 | 2 |
| **CD20** | Spark NIR 685 | BioLegend | 302366 | 2 |
| **IgD** | AF700 | BD Biosciences | 561302 | 2 |
| **CD19** | APC-Cy7 | Thermo Fisher Scientific | A15391 | 2 |
| **CD4** | cFluor R840 | Cytek | R7-20165 | 2 |
| **CD138** | BUV737 | BD Biosciences | 612834 | 2 |
| **CD21** | BUV496 | BD Biosciences | 750614 | 2 |
| **IgG3** | BUV563 | BD Biosciences | 753741 | 2 |
| **IgG1** | IgG1-BV711 | BD Biosciences | 753704 | 2 |
| **CD5** | Pacific Blue | BioLegend | 364024 | 2 |
| **CD1d** | BV480 | BD Biosciences | 746435 | 2 |
| **IgA** | Vio Green | Miltenyi Biotec | 130-113-481 | 2 |
| **CD40** | RY586 | BD Biosciences | 753194 | 2 |
| **IgG** | PE-CF594 | BD Biosciences | 562538 | 2 |
| **CD38** | PE-Fire 640 | BioLegend | 397218 | 2 |
| **CD25** | cFluor BYG710 | Cytek | R7-20585 | 2 |
| **Viability** | Zombie NIR | BioLegend | 423105 | 2 |
| **Streptavidin** | BV605 | BD Biosciences | 563260 | 3 |
| **Streptavidin** | BV786 | BD Biosciences | 563858 | 3 |
| **Streptavidin** | RB780 | BD Biosciences | 570517 | 3 |
| **Streptavidin** | Vio Bright B515 | Miltenyi Biotec | 130-108-993 | 3 |
| **Streptavidin** | BV421 | BD Biosciences | 563259 | 3 |
| **Streptavidin** | PE-Vio 770 | Miltenyi Biotec | 130-106-793 | 3 |
